# Safety and Immunogenicity of the COVID-19 Vaccine BNT162b2 in Patients Undergoing Chemotherapy for Solid Cancer

**DOI:** 10.1101/2021.10.25.21265324

**Authors:** Yohei Funakoshi, Kimikazu Yakushijin, Goh Ohji, Wataru Hojo, Hironori Sakai, Ryo Takai, Taku Nose, Shinya Ohata, Yoshiaki Nagatani, Taiji Koyama, Akihito Kitao, Meiko Nishimura, Yoshinori Imamura, Naomi Kiyota, Kenichi Harada, Yugo Tanaka, Yasuko Mori, Hironobu Minami

**Affiliations:** Division of Medical Oncology/Hematology, Department of Medicine, Kobe University Hospital and Graduate School of Medicine, Kobe, Japan; Division of Infection Disease Therapeutics, Department of Microbiology and Infectious Diseases, Kobe University Hospital and Graduate School of Medicine, Kobe, Japan; R&D, Cellspect Co., Ltd., Morioka, Japan; Division of Gastrointestinal Surgery, Department of Surgery, Kobe University Hospital and Graduate School of Medicine, Kobe, Japan; Cancer Center, Kobe University Hospital, Kobe, Japan; Division of Urology, Department of Surgery, Kobe University Hospital and Graduate School of Medicine, Kobe, Japan; Division of Thoracic Surgery, Department of Surgery, Kobe University Hospital and Graduate School of Medicine, Kobe, Japan; Division of Clinical Virology, Center for Infectious Diseases, Kobe University Graduate School of Medicine, Kobe, Japan

**Keywords:** SARS-CoV-2 vaccination, cancer patients, cytotoxic chemotherapy, immune checkpoint chemotherapy

## Abstract

**Background:** Although COVID-19 severity in cancer patients is high, the safety and immunogenicity of the BNT162b2 mRNA COVID-19 vaccine in patients undergoing chemotherapy for solid cancers in Japan have not been reported.

**Methods:** We investigated the safety and immunogenicity of BNT162b2 in 41 patients undergoing chemotherapy for solid cancers and in healthy volunteers who received 2 doses of BNT162b2. We evaluated serum IgG antibody titers for S1 protein by ELISA at pre-vaccination, prior to the second dose and 14 days after the second vaccination in 24 cancer patients undergoing cytotoxic chemotherapy (CC group), 17 cancer patients undergoing immune checkpoint inhibitor therapy (ICI group) and 12 age-matched healthy volunteers (HV group). Additionally, inflammatory cytokine levels were compared between the HV and ICI groups at pre and the next day of each vaccination.

**Results:** Anti-S1 antibody levels were significantly lower in the ICI and CC groups than in the HV group after the second dose (median optimal density: 0.241 [0.063-1.205] and 0.161 [0.07-0.857] vs 0.644 [0.259-1.498], *p* = 0.0024 and *p* < 0.0001, respectively). Adverse effect profile did not differ among the three groups, and no serious adverse event occurred. There were no differences in vaccine-induced inflammatory cytokines between the HV and ICI groups.

**Conclusion:** Although there were no significant differences in adverse events in three groups, antibody titers were significantly lower in the ICI and CC groups than in the HV group. Further protection strategies should be considered in cancer patients undergoing CC or ICI.

**Mini abstract:** Titers of anti-S1 antibody after the second dose of BNT162b2 were significantly lower in patients with solid tumors undergoing active anticancer treatment than in the healthy volunteers.

## Introduction

Compared with individuals with the corona virus disease 2019 (COVID-19) without cancer, individuals with COVID-19 and cancer have been characterized as having severe outcomes and mortality. (1, 2) In a meta-analysis of 52 studies involving 18,650 patients with COVID-19 and cancer, the incidence of mortality was high at 25.6%. (3) Additionally, some studies raised concerns about the delayed diagnosis of primary and/or recurrent cancers and delayed treatment due to COVID-19. (4) Accordingly, experts have generally recommended that patients with cancer should receive vaccines to protect against COVID-19.

The BNT162b2 mRNA COVID-19 vaccine that leads to transient expression of the SARS-CoV-2 spike protein was developed in 2020. (5) The phase 3 trial of the vaccine showed 95% efficacy in preventing symptomatic COVID-19 infection. (6) However, data concerning the efficacy and safety of this vaccine for cancer patients was insufficient, because they were excluded from the initial registration trials. (6)

Most recently, several studies have reported that SARS-CoV-2 antibody titers after vaccination were significantly lower in patients with solid tumors undergoing chemotherapy than in healthy volunteers. (7-10) Interestingly, the efficacy of COVID-19 vaccines may vary by race in view of the variation in the major histocompatibility complex molecules. (11) Further, the Review Report of BNT162b2 by the Pharmaceuticals and Medical Devices Agency (PMDA) of Japan revealed that vaccine efficacy in Asian populations might be lower than in the total population (74.6 vs 95.0%). (12) Considering these results, data collection in different regions and races is important. However, no data on the safety and immunogenicity of the COVID-19 vaccines for cancer patients undergoing chemotherapy have been reported in Japan. Here, we investigated the safety and immunogenicity of BNT162b2 in Japanese patients with solid tumors receiving cytotoxic chemotherapy or immune-checkpoint inhibitor (ICI) therapy. Additionally, in the assessment of adverse reactions, we hypothesized that ICI-activated immunity might induce excessive inflammatory effects after BNT162b2. To test this, levels of various inflammatory cytokines before and the day after each vaccination were compared between the HV and ICI groups.

## Patients and Methods

### Study Design

Patients with solid tumors (histologically diagnosed) undergoing active anticancer treatment (cytotoxic chemotherapy or ICI therapy) at Kobe University Hospital between May 2021 and August 2021 were enrolled, together with age-matched healthy volunteers. All participants were vaccinated with 2 doses of BNT162b2. Peripheral blood samples were collected at pre-vaccination (within 7 days prior to the first dose), within 3 days prior to the second dose and 14 days (+/- 7 days) after the second dose of BNT162b2. For cytokine assay, additional blood samples were collected at the next day of each vaccination in the HV and ICI groups. Exclusion criteria included a documented COVID-19 infection (positive PCR test result) within 14 days after the second dose. Vaccine-related adverse events were evaluated by Common Terminology Criteria for Adverse Events 5.0, except for fever, which we defined as Grade 1 37.5 – 37.9°C, Grade 2 38.0 – 38.9°C, Grade 3 39.0 – 39.9°C and Grade 4 >40.0°C in the axilla. The study was approved by the Kobe University Hospital Ethics Committee (No. B2056714) and was conducted in accordance with the Declaration of Helsinki. All patients provided written informed consent for this study.

### Sample collection and measurement of antibody titers against S1 protein

Serum samples were obtained by centrifuging the blood samples for 10 min at 1000 × *g* at room temperature, and were immediately transferred to a freezer kept at - 80°C.

Antibody titers against S1 were measured using the QuaResearch COVID-19 Human IgM IgG ELISA kit (Spike Protein-S1) (Cellspect, Inc., RCOEL961S1, Iwate, Japan). This kit is based on the indirect ELISA method, and comes with different immobilized antigenic proteins. The plate of the ELISA kit (Spike Protein-S1) is immobilized with a recombinant spike protein (S1, 251-660AA) of SARS-CoV-2 expressed in *Escherichia coli*. Serum samples were diluted 1:200 in 1% BSA/PBST for RCOEL961S1.The plates were read at 450 nm with an SH-1200 plate reader (Corona Electric Co. Ltd.) in accordance with the manufacturer’s measurement protocol.

### Measurement of inflammatory cytokines

Serum inflammatory cytokines (Interferon [IL]-2, IL-4, IL-6, IL-8, IL-10, tumor necrosis factor [TNF] -α, interferon gamma [IFN]-γ, and granulocyte macrophage-colony stimulating factor [GM-CSF]) were probed using Bio Plex Pro Human Cytokine plex Panel (Bio-Rad, CA, USA) multiplex magnetic bead-based antibody detection kits following the manufacturer’s instructions. Standard curves for each analyte were generated using standards provided by the manufacturer and the collected data were analyzed using Bio-Plex Manager™ Software version 6.1 (Bio-Rad). All assays were performed by Filgen (Tokyo, Japan).

## Results

### Patients

Forty-one Japanese patients with solid tumors and 12 healthy volunteers were enrolled in this study. Among the 41 patients, 24 were undergoing cytotoxic chemotherapy (CC group) and 17 patients were undergoing ICI therapy (ICI group). Treatments are shown in Table 1. Among the ICI group, 14 patients were previously treated with cytotoxic chemotherapy with or without radiation therapy and two patients were treated with radiation therapy alone before ICI therapy (Table 1). Both groups received the first and second doses of BNT162b2 during the course of these therapies.

**Table 1.** Patient characteristics.

|  | HV group | CC group | ICI group |
| --- | --- | --- | --- |
| **N (Female, Male)** | 12 (7, 5) | 24 (8, 16) | 17 (4, 13) |
| **Median age (range)** | 76.5 (67–82) | 72.5 (66–82) | 75 (64–84) |
| **Solid malignancies** |  |  |  |
| Head and neck | - | 2 | 3 |
| Renal | - | 0 | 1 |
| Bladder/Renal Pelvis and Ureter | - | 0 | 3 |
| Esophageal | - | 0 | 3 |
| Stomach | - | 2 | 1 |
| Colorectal | - | 10 |  |
| Melanoma | - | 0 | 2 |
| Pancreas | - | 8 | 0 |
| Bile duct | - | 0 | 1 |
| Others | - | 2 | 3 |
| **CC therapy** |  |  |  |
| Gem + nab-PTX | - | 7 | - |
| FOLFOX-based | - | 3 | - |
| CapeOX-based | - | 3 | - |
| PTX (with or without cetuximab) | - | 3 | - |
| SOX-based | - | 2 | - |
| CPT-11 (with or without ramucirumab) | - | 2 | - |
| Others | - | 4 | - |
| **ICI therapy** |  |  |  |
| Nivolumab | - | - | 10 |
| Pembrolizumab | - | - | 5 |
| Avelumab + Axitinib | - | - | 1 |
| Atezolizumab + Bevacizumab | - | - | 1 |
| **Anti-cancer therapy before ICI** |  |  |  |
| CC or CC + RT | - | - | 14 |
| RT alone | - | - | 2 |
| none | - | - | 1 |

HV; healthy volunteers, CC; cytotoxic chemotherapy, ICI; immune checkpoint inhibitor therapy, Gem; gemcitabine, nab-PTX; albumin-bound paclitaxel, CPT-11; irinotecan, FOLFOX; oxaliplatin and fluorouracil / folinic acid, CapeOX; oxaliplatin and capecitabine, SOX; oxaliplatin and tegafur-gimeracil-oteracil potassium, RT; radiation therapy,

### Serological outcomes

Antibody titers after the second dose were significantly higher than those at pre-vaccination in all groups (Figure 1A). However, anti-S1 antibody levels in both the ICI and CC groups were significantly lower than those in the HV group after the second dose (median optimal density: 0.241 [0.063-1.205] and 0.161 [0.07-0.857] vs 0.644 [0.259-1.498]; *p* = 0.0024 and *p* < 0.0001, respectively) (Figure 1A and B).

**Figure 1.**
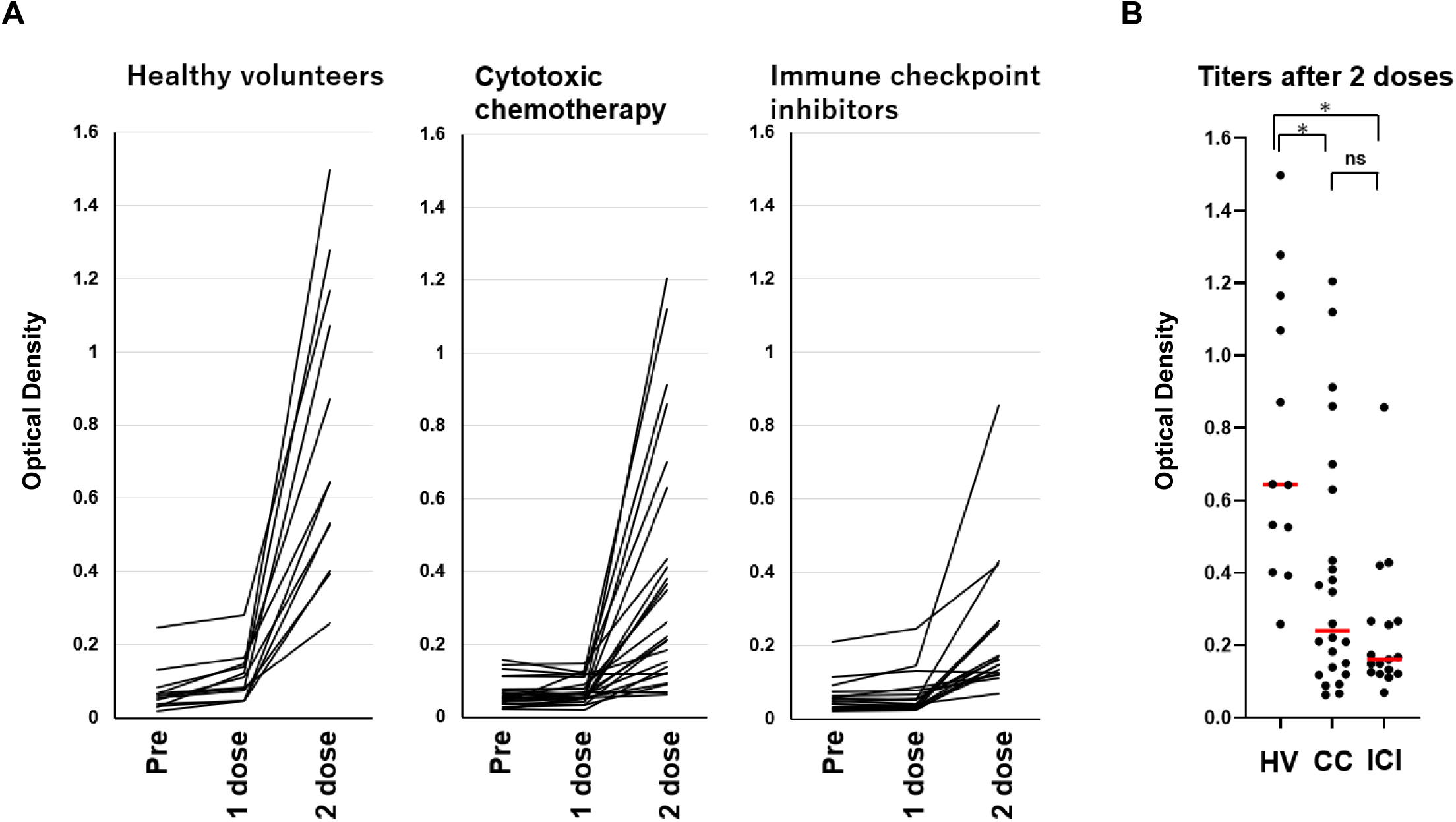
**(A)** Humoral quantitative anti-spike 1 (S1) antibody response at pre-vaccination (within 7 days prior to the first dose), within 3 days prior to the second dose and 14 days (+/- 7 days) after the second dose of BNT162b2 mRNA SARS-CoV-2 vaccine in healthy volunteers (HV group) (n = 12), patients treated with cytotoxic chemotherapy (CC group) (n = 24) and patients treated with immune checkpoint inhibitors (ICI group) (n = 17). (**B)** S1 antibody titers 14 days (+/- 7 days) after the second vaccination dose in the HV, CC and ICI groups. The red lines indicate the median value of optical density in each group. ns; not significant. *, *p* < 0.01.

### Adverse events and cytokine profile

There was no clinical difference in the profile of vaccine-related adverse events among the three groups (Figure 2), and no serious adverse event (> Grade 3) was reported in any participant.

**Figure 2.**
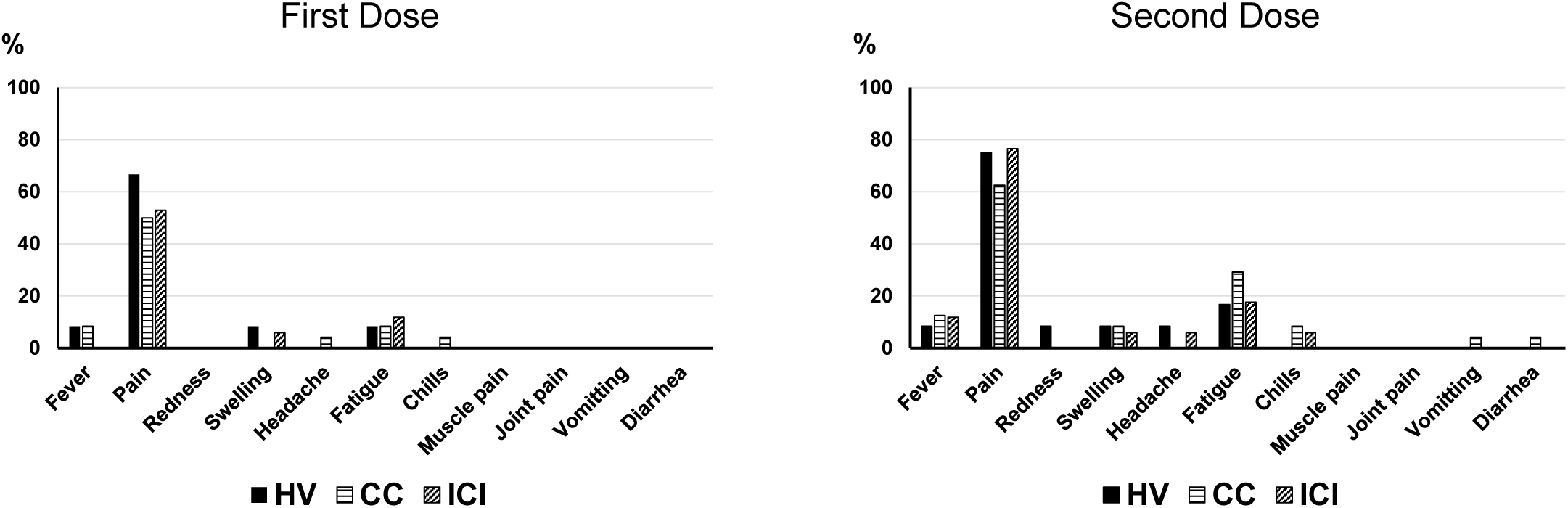
Vaccine-related adverse events reported within 7 days after first and second dose of BNT162b2 in healthy volunteers (HV group) (n = 12), and patients treated with cytotoxic chemotherapy (CC group) (n = 24) and immune check point inhibitors (ICI group) (n = 17).

The impact of vaccination on ICI-activated immunity was investigated by measuring cytokine levels before and the day after each vaccination. However, levels of inflammatory cytokines did not show any significant changes in the HV or ICI groups (Figure 3). Results showed no meaningful differences between the two groups.

**Figure 3.**
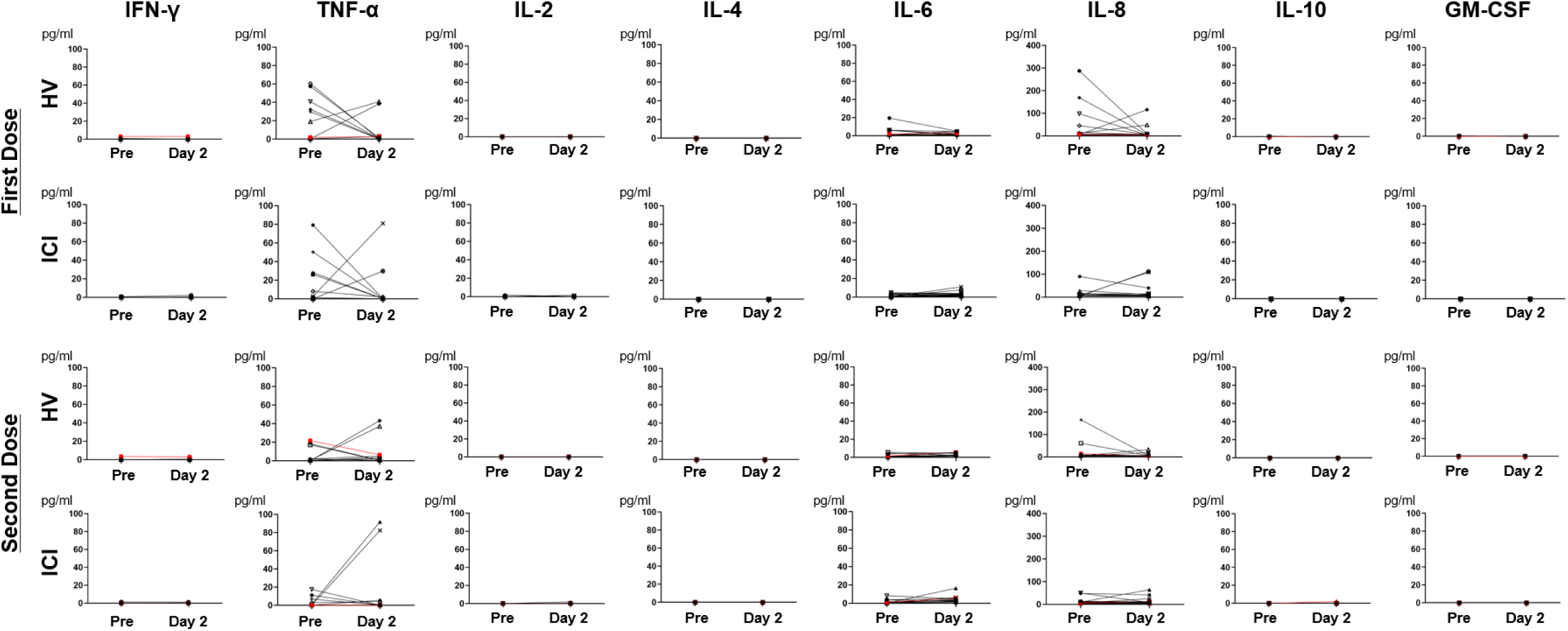
Change in serum levels of interleukin (IL)-2, IL-4, IL-6, IL-8, IL-10, interferon (IFN)-γ, tumor necrosis factor (TNF)-α and granulocyte macrophage-colony stimulating factor (GM-CSF) in healthy volunteers (HV group), and patients treated with immune check point inhibitors (ICI group). Days indicate the day after each vaccination. The red lines indicate participants with fever.

## Discussion

Because cytotoxic chemotherapy affects bone marrow and suppresses the immune system, concern has been raised that these drugs impair the efficacy of SARS-CoV-2 vaccines in triggering the humoral immune response. Most recently, several study groups reported that antibody titers of patients with cancer who were receiving active systemic chemotherapy were significantly lower than those of healthy volunteers. (7-9) Consistent with these reports, our Japanese data also indicated that anti-S1 antibody levels in cancer patients treated with active chemotherapy were significantly lower than those observed in the HV group after the second dose. Interestingly, anti-S1 antibody levels were significantly lower in not only the CC group but also in the ICI group. Given that the ICI enhances the immune system, this result was unexpected. Another research group reported similar results in a small number of cases (n = 14). (7) It is possible that the effect of cytotoxic chemotherapy and/or RT before ICI therapy and the immunosuppressed state due to malignancy itself suppress antibody production.

Although the presence of lower titers than in healthy volunteers does not prove that humoral immunity has no preventive effect, further protection strategies should be considered in these cancer patients. To protect these patients from COVID-19, we first recommend that they continue to wear a face mask and wash their hands even after vaccination. (13, 14) Second, a third dose (booster dose) of the COVID-19 vaccines might be effective. The efficacy of three doses has been reported in patients with solid organ transplantation who also have impaired antibody production due to immunosuppressants. (15)

Inflammatory reactions including fever have been reported as major adverse reactions after vaccination with mRNA-containing lipid nanoparticles (LNPs) such as BNT162b2. A study group has reported that vaccines based on mRNA-containing LNPs induced high production of inflammatory cytokines (IL-6, GM-CSF and IL-1β) in mice 24 hours after vaccination. (16) Because we were concerned that ICI-activated immunity might induce excessive inflammatory effects, inflammatory cytokine levels were measured before and after vaccination (Day 2) and compared between the HV and ICI groups. Contrary to the results in the mouse model, there was no trend toward an increase in levels of inflammatory cytokines after vaccination in either the HV group or ICI group, despite our consideration that the immune systems of these patients were activated. Further, cytokine level was not increased even in patients who had fever after vaccination. Overall, levels of all measured cytokines were low. Consistent with these results, we found no clinical differences in the profile of adverse reactions among the three groups, and no serious adverse event was reported in any participant.

In conclusion, although there was no significant difference in adverse events in the three groups, antibody titers in Japanese patients treated with cytotoxic chemotherapy and ICI were significantly lower than those in the HV group. Further protection strategies should be considered.

## Data Availability

All data produced in the present study are available upon reasonable request to the authors.

## Conflicts of Interest

Hironobu Minami has received research grants and honoraria from Pfizer. Wataru Hojo and Hironori Sakai are employed by Cellspect Co., Ltd. Kimikazu Yakushijin has received honoraria from Pfizer. The other authors declare no potential conflicts of interest.

## Notes

### Funding Statement

This study was funded by Division of Medical Oncology/Hematology, Department of Medicine, Kobe University Hospital and Graduate School of Medicine.

### Author Declarations

The study was approved by the Kobe University Hospital Ethics Committee (No. B2056714) and was conducted in accordance with the Declaration of Helsinki. All patients provided written informed consent for this study.

